# The impact of genetic and sociodemographic factors on life course trajectories of physical-mental health multimorbidity in a UK South Asian cohort

**DOI:** 10.1101/2025.10.13.25337676

**Authors:** Daniel Stow, Ruby Tsang, Ioanna Katzourou, Qinqin Huang, Miriam Samuel, The Genes & Health Team, The LIfespaN multimorbidity research Collaborative (LINC), Rupert Payne, Peter Holmans, Nic Timpson, Inês Barroso, Hilary Martin, Jack F. G. Underwood, James T.R. Walters, Megan L. Wood, Marianne B.M. van den Bree, Rohini Mathur, Sarah Finer

## Abstract

**Background:** UK South Asian populations have a high risk of many cardiometabolic and internalising mental health conditions such as depression and anxiety, and they commonly co-exist as physical and mental health multimorbidity. However, the emergence of this multimorbidity across the lifecourse, and its underlying aetiology, is rarely studied.

**Methods:** We studied Internalising and Cardiometabolic MultiMorbidity (ICM-MM) in a longitudinal cohort with linked genetic and health data from Genes & Health. ICM-MM was defined as lifetime occurrence of ≥ 1 internalising condition AND ≥ 1 cardiometabolic condition. Using multi state models, we investigated ICM-MM trajectories and their effect on risk of cardiovascular event (CVE) or death, accounting for competing risks. We used flexible parametric models to estimate baseline hazards for transitions between health states, adjusting for gender, age, ethnicity, deprivation, smoking, and a novel polygenic risk score for ICM-MM (ICM-MM_PRS_)

**Findings:** In our cohort of 25,641 British Bangladeshi and Pakistani people (median follow-up 10.5 years, median age 31.1 years at cohort entry), women had higher risk of internalising conditions and subsequent ICM-MM, whilst men were at higher risk of CVE. Younger age was associated with higher risk of internalising conditions. Bangladeshi ethnicity, deprivation, and smoking were all associated with higher ICM-MM risk. 10-year CVE risk was higher for transitioning from cardiometabolic morbidity to ICM-MM in midlife than internalising morbidity to ICM-MM. ICM-MM_PRS_ was associated with higher risk of transitioning from cardiometabolic morbidity to ICM-MM than internalising morbidity to ICM-MM.

**Interpretation:** The burden of multimorbidity is high in the population and develops at an early age. Young Bangladeshi women are at highest risk of ICM-MM, while men are at higher risk of CVE. Detection and intervention strategies for physical-mental health multimorbidity targeted early in the lifecourse, for those at risk.

**Funding**: MRC[MR/W014416/1]

## Introduction

Over a third of the world’s population live with multimorbidity^1^ - the co-occurrence^2^ or lifetime occurrence^3^ of two or more long-term health conditions. Multimorbidity has a profound impact on the lives of patients and their families, and has led to increasing demands on health and social care systems.^4^ Multimorbidity disproportionately affects those living in deprived circumstances,^5^ and the prevalence of multimorbidity is projected to rise as populations age, ^6^ further increasing existing global health inequalities.

Numerous studies have identified patterns of frequently co-occurring long-term conditions - or multimorbidity clusters - highlighting prevalent combinations,^7^ their early onset in south Asian populations in the UK, and their impact on health service utilisation and cost.^8–10^ However, multimorbidity remains under-studied despite its major impact at individual and population level and on health services. There is an urgent need to study how and why multimorbidity develops so that effective prevention and intervention strategies can be evaluated and implemented.^3^ Some research is starting to address these knowledge gaps by identifying the sequences of conditions or ‘trajectories’ leading to multimorbidity. These studies have identified those trajectories that are most detrimental to reduced life-expectancy^11^ and have also highlighted the association of multimorbidity trajectories with age, gender, socio-economic position.^12,13^ However, this prior research does not elucidate aetiological mechanisms underlying the development of multimorbidity trajectories and they use, if at all, broad ethnicity categories (e.g. south Asian) that are confounded by socio-economic position and do not reflect the social, behavioural and ancestral differences within them.

In the UK, Pakistani and Bangladeshi populations are more likely than other south Asian and European populations to live with multiple long-term conditions and report poorer health. ^14,15^ This is supported by epidemiological and genomic literature demonstrating the earlier onset, and greater genetic risk of conditions such as type 2 diabetes,^16^ which are a common component of multimorbidity clusters. There is emerging literature suggesting that commonly co-occurring conditions such as depression and type 2 diabetes may share some common aetiology through immunological and genetic pathways.^17,18^ However, whilst the genetic basis of single common conditions is well -characterised and leading to combined genetic and clinical risk models being implemented within health systems such as the NHS,^19,20^ there is very little understanding of the shared genetic risk of multimorbidity, and how this manifests across the lifecourse. Without such knowledge, the prediction and prevention of multimorbidity may be limited to existing clinical risk tools (particularly for single conditions), which may not offer effective screening for multimorbidity risk.

We sought to characterise health trajectories in a multimorbidity cluster composed of internalising mental-health and cardiometabolic conditions in a large UK sample of British Bangladeshi and Pakistani South Asian individuals and investigate, for the first time, whether a multimorbidity polygenic risk tool alongside traditional risk factors would identify those at risk. We selected this population due to the greater burden and earlier onset of several of the single conditions in this multimorbidity cluster, and to redress the lack of representation of south Asian populations in genomic research.^21^

We focused on a cluster of physical and mental health multimorbidity comprising internalising^23^ mental-health conditions and cardiometabolic conditions: one of the most common types of physical and mental health multimorbidity in older age.^24^ This constellation of conditions, or similar, are commonly observed in clustering studies,^1^ ^25^ and account for a significant cost to health systems.^26^ and yet there is a lack of evidence on how to prevent, or manage them effectively in multimorbidity care models.^27,28^

We used multi-source electronic health records to describe the transitions from a healthy state to the development of either an internalising or cardiometabolic condition and subsequent multimorbidity, followed by a cardiovascular event and death from any of these conditions. We then investigated the influence of ethnicity, sociodemographic and genetic risk, using a novel polygenic risk score for multimorbidity, on the progression through these trajectories.

## Methods

### Patient and public involvement

Members of the public with lived experiences of multiple long-term conditions contributed to the design of the study - highlighting priority areas and informing the research questions - and are acknowledged in the acknowledgements section.

### Ethics

Genes & Health was approved by the National Research Ethics Committee (London and Southeast), and the Health Research Authority (reference 13/LO/124). Written informed consent allowing the collection, analysis, and publication of results from health and genetic data is obtained from all study volunteers.

### Data sources and setting

Genes & Health (G&H) is an ongoing, population-based study that recruits participants of British Pakistani and British Bangladeshi ethnicity aged 16 and over from East London, Manchester, Birmingham, and Bradford in the United Kingdom. At recruitment, participants complete a brief demographic questionnaire, consent to secure linkage to their National Health Service (NHS) electronic health records, and provide a saliva sample for DNA studies.^29^

Health phenotypes in G&H are captured from primary care providers across the inner north-east London boroughs of City & Hackney, Newham, Tower Hamlets & Waltham Forest).^30^ Linkage to NHS Digital datasets provides Hospital Episode Statistics (HES) Admitted Patient Care (APC) records from hospitals in England and Wales. We accessed information on underlying cause and date of death from linked Office of National Statistics (ONS) Civil Registrations data. All health phenotypes were defined using existing SNOMED (primary care data) and ICD-10 (HES-APC) publicly available codelist resources.^31^ Saliva sample genotyping was performed with the Illumina Infinium Global Screening Array V3 Chip and imputation via TOPMed servers^32^ using the r3 reference panel.^33^

### Cohort design

We generated a longitudinal cohort nested in G&H using routine electronic health record linkages. The start of the follow up period was defined as the most recent of, i) date of registration with a GP practice contributing primary care data to G&H, ii) date the individual turned 16 years of age (minimum eligible age of G&H volunteers), or iii) date of HES-APC linkage initiation (1st April 1997 per Herbet et al., 2017).^34^ The end of the follow up period was defined as the earliest of i) date of loss to follow up (12 months after the most recent primary care or HES-APC record),^35^ ii) date of death, taken from Office of National Statistics (ONS) civil registration mortality data, iii) date of administrative censoring (earliest of data refresh from primary care or HES data [set to November 2024]), or iv) date the participant de-registered from a GP practice contributing data to G&H.

### Participants

Eligible participants were age ≥16 years old, with ≥1 record in the G&H primary care dataset, no record of any internalising or cardiometabolic conditions before start of follow-up, and total follow-up time >0 days.

### Multimorbidity definition

This work is part of the LIfespaN multimorbidity research Collaborative (LINC) programme.^36^ LINC defines Internalising and CardioMetabolic MultiMorbidity (ICM-MM) as the lifetime occurrence of ≥1 internalising mental health condition (depression, anxiety, somatoform disorders) AND ≥one cardiometabolic condition (hypertension, type 2 diabetes [T2D], obesity, dyslipidaemia, chronic kidney disease [CKD]) for further information see <u>SUPPLEMENTAL A</u>). Age of onset was defined as the first occurrence of any relevant code in primary care or HES records.

### Outcome measures

The primary endpoint for this study was a cardiovascular event (CVE),^37^ one of the leading causes of death globally, and a common consequence of the cardiometabolic conditions in our ICM-MM definition.^38^ CVE was defined as the earliest occurrence in the electronic healthcare record data of, or where the underlying cause of death in ONS data^39^ was due to: atrial fibrillation and flutter (AF), heart failure (HF), coronary heart disease (CHD), peripheral vascular disease (PVD), cerebrovascular disease (TIA/stroke), and end stage renal disease (ESRD). All other underlying causes of mortality (death) were grouped together as a competing risk state in the multi-state model (<u>FIGURE1</u>).

**FIG 1.**
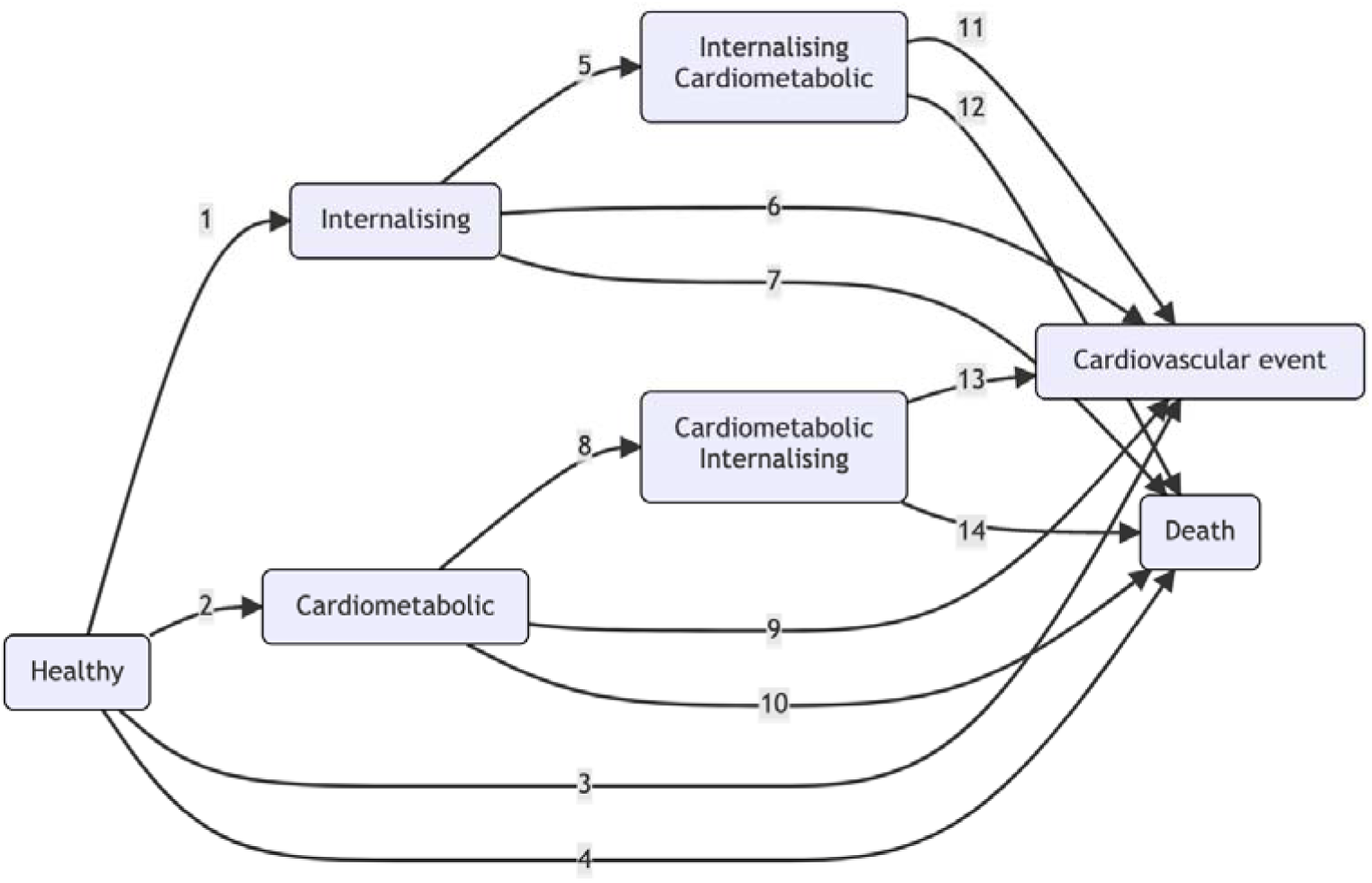
Multi state model

### Exposure measures

#### Demographic variables

Participants’ self-reported gender, and ethnicity were taken from the G&H demographic questionnaire. Participants responding ‘other’ to the ethnicity item were excluded from this analysis. Date of birth was taken from participants’ primary care electronic records. We used Index of Multiple Deprivation (IMD 2019)^40^ quintiles from primary care records, collapsing the top three quintiles (1 = most deprived… 3+ = least deprived) due to the small number of participants living in the least deprived areas and to aid model convergence. Smoking status was defined as ‘ever’ or ‘never’ at study baseline using existing codelists.^41^

#### Genetic risk

We have previously described the methods used to derive a novel PRS for ICM-MM: ICM-MM_PRS_. Briefly, we created seven trait specific polygenic risk scores (PRS_TRAIT_) for anxiety, depression, T2D, pulse pressure (a proxy for hypertension), body mass index ([BMI]a proxy for obesity), chronic kidney disease (CKD), low-density lipoprotein cholesterol ([LDL-C]a proxy for dyslipidaemia) in UK BioBank,^42^ using PRS-CS^43^ and summary statistics from published GWAS excluding UKB. We used elastic net regression, implemented in the R package ‘PRSmix’,^44^ to test the optimal weighting and linear combination of the seven PRS_TRAIT_ to predict ICM-MM in UK BioBank participants. The final ICM-MM_PRS_ combined PRS_TRAIT_ for (in order of weighting, high to low) depression, pulse pressure, T2D, BMI and LDL-C.

For this analysis, we used the SNP effect weights from PRS-CS to calculate PRS_TRAIT_ for all individuals in G&H using PLINK 2.0, and the PRS weights from PRSMix to calculate ICM-MM_PRS_ in G&H. To aid interpretation in our results we regressed out the first 20 genetic principal components and then z-transformed ICM-MM_PRS_.

We carried out a supplemental sensitivity analysis following the same process described inbut for over 600 PRS from the PGS catalogue. For this analysis we used a 50% random subsample of G&H data for the training the Elastic Net, and the remaining 50% hold out set for testing the resulting PRS as an exposure variable in the multi-state models. [SUPPLEMENTAL B]

## Statistical analysis

We used a semi-Markov (‘clock-reset’) multi state model^45^ to describe fourteen transitions between seven health states (<u>FIGURE1</u>), starting from ‘otherwise healthy’ - free from any ICM-MM conditions at or before study baseline. Transitions were allowed from ‘otherwise healthy’ to single disease states (any internalising OR any cardiometabolic condition) and then to ICM-MM, accounting for disease ordering (i.e. internalising → cardiometabolic versus cardiometabolic → internalising), and then to CVE or death from either ICM-MM trajectory. Direct transitions to CVE and death were allowed from any other health state in the model (e.g. otherwise healthy → death, internalising → CVE), but transitions from CVE or death to other health states were not (i.e. these were ‘absorbing states’).

Baseline hazards for each transition were estimated separately using restricted cubic splines as described in Royston and Parmar, 2002^46^ using ‘flexsurvspline’ from the ‘flexsurv’ package.^47^ We selected the number of interior knots for each model via Akaike information criterion (AIC) and comparison of modelled vs observed survival plots for each transition. All models were adjusted for baseline age, IMD, gender, ethnicity, smoking status, and ICM-MM_PRS_. Baseline age was defined as the age at entry into each state separately (i.e. baseline age for the ‘otherwise healthy’ state is equal to the age at cohort entry and increases for each subsequent change in health state). Participants with missing covariates were dropped (i.e. a complete case analysis).

We assessed the proportionality assumption via Schoenfeld residuals plotted against transformed time. Where we observed non-proportional hazards, we allowed spline parameters to vary with the relevant covariate values (i.e. time-varying effects). For full details of model selection and final model specifications, please see [SUPPLEMENTAL C]

### State occupation probabilities

Predicted probabilities of being in state_i_ at time_t_ were simulated using 10,000 individuals for 100 evenly spaced time points over a ten-year window using the sim.fmsm command in the ‘flexsurv’ package.^47^ All simulations were for a pre-specified covariate pattern: a Bangladeshi woman living in IMD quintile 1 (most deprived) with ICM-MM_PRS_ z-score of 0 (‘average risk’), and a ‘never’ smoker age 40 at the start of follow-up, chosen due to its relevance and generalisability to the NHS Health Check which is offered from age 40 in England and Wales.^48^ We also examined the impact of varying age at start of follow up, retaining all other reference covariate patterns for comparison.

### Condition ordering and CVE risk

To test the impact of multimorbidity on future risk of CVE, we simulated state occupation probabilities for all transitions in the multi state model and then contrasted single condition trajectories with the multimorbid equivalent e.g. *P(CVE* | *cardiometabolic)* versus *P(CVE | cardiometabolic* → *internalising)*. To test the impact of condition ordering in multimorbid trajectories on future risk of CVE, we simulated state occupation probabilities for all transitions in the multi state model, then contrasted *P(CVE | cardiometabolic* → *internalising)* with *P(CVE | internalising* → *cardiometabolic)* over a range of ages of ICM-MM onset.

### Sociodemographic and genetic contrasts

To test the impact of sociodemographic factors and ICM-MM_PRS_ on state occupation probabilities, we generated contrasts between the reference covariate pattern, and the contrast of interest (e.g. *P(state_i_ time_t_ | female) - P(state_i_ time_t_ | male*). Confidence intervals for contrasts were generated using bootci.fmsm in flexsurv,^47^ with n=400 iterations.

## RESULTS

### Demographics

The flow diagram for cohort eligibility is shown in [<u>FIGURE2</u>]: 25,641G&H participants were included in the complete case analysis;13,995 (54.6) were female, and 11,646 (45.4) male. Baseline characteristics for the cohort are shown in [TABLE1]. Median age at cohort entry was 31.1 years (IQR: 24.1–38.3), median follow up time was 10.5years (IQR: 6.4–16.8), and loss to follow up (2.1%) was acceptable.^49^

**FIG 2.**
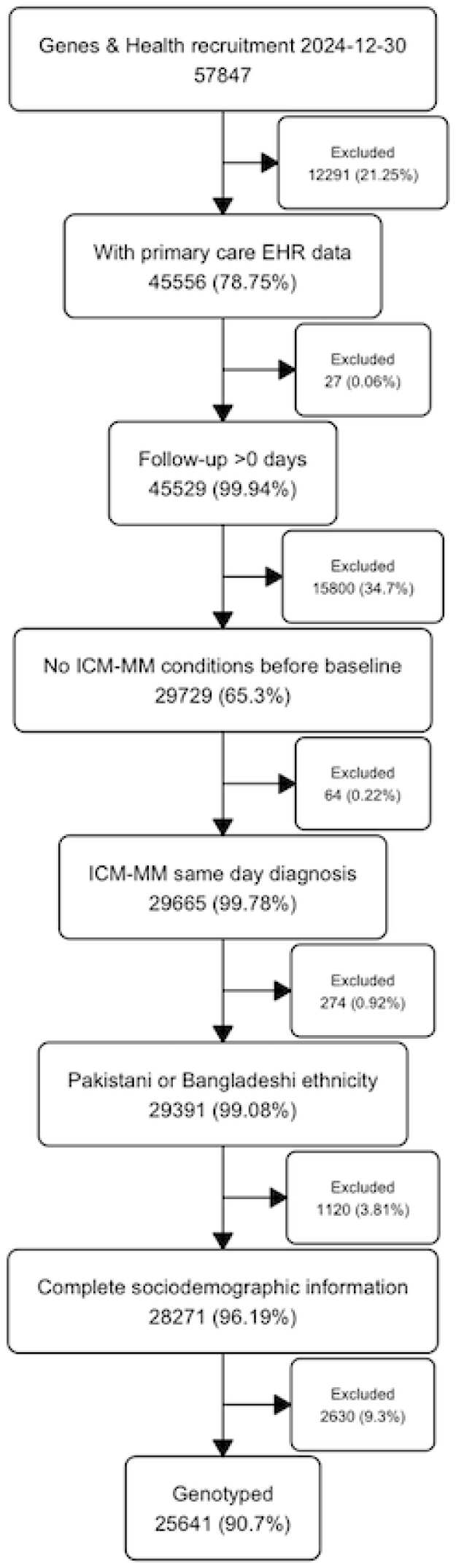
Study flow chart

**TABLE 1.** Study participants

|  | Bangladeshi |  | Pakistani |  | Overall |  |
| --- | --- | --- | --- | --- | --- | --- |
|  | Female<br>(N=9217) | Male<br>(N=7711) | Female<br>(N=4778) | Male<br>(N=3935) | Female<br>(N=13995) | Male<br>(N=11646) |
| **ICMM PGS** |  |  |  |  |  |  |
| Mean (SD) | -0.0146 (0.981) | -0.0597 (0.993) | -0.0405 (1.01) | -0.0660 (1.00) | -0.0234 (0.992) | -0.0619 (0.997) |
| Median [Min, Max] | -0.0225 [-3.47, 4.02] | -0.0517 [-3.87, 3.28] | -0.0485 [-3.68, 4.23] | -0.0772 [-3.58, 3.36] | -0.0281 [-3.68, 4.23] | -0.0595 [-3.87, 3.36] |
| **Age at cohort entry** |  |  |  |  |  |  |
| Mean (SD) | 29.9 (9.44) | 33.5 (10.9) | 31.0 (10.5) | 33.3 (12.0) | 30.3 (9.81) | 33.4 (11.3) |
| Median [Min, Max] | 29.2 [16.0, 80.0] | 33.8 [16.0, 87.9] | 29.9 [16.0, 84.4] | 32.6 [16.0, 82.0] | 29.4 [16.0, 84.4] | 33.4 [16.0, 87.9] |
| **IMD Quintile \*** |  |  |  |  |  |  |
| 1 | 3122 (33.9%) | 2566 (33.3%) | 708 (14.8%) | 607 (15.4%) | 3830 (27.4%) | 3173 (27.2%) |
| 2 | 4799 (52.1%) | 4049 (52.5%) | 2818 (59.0%) | 2287 (58.1%) | 7617 (54.4%) | 6336 (54.4%) |
| 3+ | 1296 (14.1%) | 1096 (14.2%) | 1252 (26.2%) | 1041 (26.5%) | 2548 (18.2%) | 2137 (18.3%) |
| **Smoking status** |  |  |  |  |  |  |
| Never | 8075 (87.6%) | 3466 (44.9%) | 4315 (90.3%) | 2391 (60.8%) | 12390 (88.5%) | 5857 (50.3%) |
| Ever | 1142 (12.4%) | 4245 (55.1%) | 463 (9.7%) | 1544 (39.2%) | 1605 (11.5%) | 5789 (49.7%) |
| **Follow up (years)** |  |  |  |  |  |  |
| Mean (SD) | 12.2 (7.31) | 11.2 (6.89) | 13.1 (7.69) | 12.2 (7.52) | 12.5 (7.46) | 11.5 (7.13) |
| Median [Min, Max] | 10.6 [0.00273, 27.6] | 9.70 [0.00274, 27.6] | 11.4 [0.00273, 27.6] | 10.7 [0.0110, 27.6] | 10.8 [0.00273, 27.6] | 10.1 [0.00274, 27.6] |
| **Internalising** |  |  |  |  |  |  |
| Yes | 2996 (32.5%) | 1483 (19.2%) | 1509 (31.6%) | 685 (17.4%) | 4505 (32.2%) | 2168 (18.6%) |
| No | 6221 (67.5%) | 6228 (80.8%) | 3269 (68.4%) | 3250 (82.6%) | 9490 (67.8%) | 9478 (81.4%) |
| **Cardiometabolic disease** |  |  |  |  |  |  |
| Yes | 4097 (44.5%) | 3511 (45.5%) | 2134 (44.7%) | 1665 (42.3%) | 6231 (44.5%) | 5176 (44.4%) |
| No | 5120 (55.5%) | 4200 (54.5%) | 2644 (55.3%) | 2270 (57.7%) | 7764 (55.5%) | 6470 (55.6%) |
| **CMD->INT** |  |  |  |  |  |  |
| Yes | 740 (8.0%) | 423 (5.5%) | 363 (7.6%) | 206 (5.2%) | 1103 (7.9%) | 629 (5.4%) |
| No | 8477 (92.0%) | 7288 (94.5%) | 4415 (92.4%) | 3729 (94.8%) | 12892 (92.1%) | 11017 (94.6%) |
| **INT->CMD** |  |  |  |  |  |  |
| Yes | 876 (9.5%) | 374 (4.9%) | 447 (9.4%) | 174 (4.4%) | 1323 (9.5%) | 548 (4.7%) |
| No | 8341 (90.5%) | 7337 (95.1%) | 4331 (90.6%) | 3761 (95.6%) | 12672 (90.5%) | 11098 (95.3%) |
| **Cardiovascular event\*\*** |  |  |  |  |  |  |
| Yes | 357 (3.9%) | 740 (9.6%) | 270 (5.7%) | 421 (10.7%) | 627 (4.5%) | 1161 (10.0%) |
| No | 8860 (96.1%) | 6971 (90.4%) | 4508 (94.3%) | 3514 (89.3%) | 13368 (95.5%) | 10485 (90.0%) |
| **Died\*\*\*** |  |  |  |  |  |  |
| Yes | 35 (0.4%) | 40 (0.5%) | 21 (0.4%) | 23 (0.6%) | 56 (0.4%) | 63 (0.5%) |
| No | 9182 (99.6%) | 7671 (99.5%) | 4757 (99.6%) | 3912 (99.4%) | 13939 (99.6%) | 11583 (99.5%) |

\*Lowest quintile is most deprived \*\*Including deaths due to CVE \*\*\*All cause mortality (ONS civil registrations)

During follow up, n=6,673 (26.0), participants developed internalising conditions (median age 35.8 years, IQR: 28.7-43.6, n= 11,407 (44.5) developed cardiometabolic conditions (median age 41.6, IQR: 35.6-48.3), and n=3,603 (14.1) developed ICM-MM (median age 42.9, IQR: 36.3–50.3). Cardiovascular event (CVE) was experienced by n=1,788 (7.0) participants (median age 52.7.9, IQR:45.1-61.0) and n=119 (0.5%) participants died due to all (non-CVE) causes (median age 57.5, IQR:47.8–71.2).

### Multi state models

#### Sequence and number of events

In total, 1,871 (37.87%) of participants with an internalising condition went on to develop a cardiometabolic condition, median age 41.5 years (IQR 35.4 – 48.2) and 1,732 (18.16%) of people with a cardiometabolic condition went on to develop an internalising condition, median age 44.7 years (IQR 37.4 – 53.3),

#### State occupation probabilities

[<u>FIGURE4a</u>] shows the state occupation probabilities adjusted for the reference covariate pattern and for five additional baseline ages ([<u>FIGURE4b</u>] shows the same plot for males). These projections show how the probability of developing internalising and cardiometabolic conditions, and ICM-MM, increases over time, and also increases with baseline age. The burden of morbidity in this cohort is high - notably, for a 40-year-old Bangladeshi woman (or man) living in the most deprived areas, the probability of remaining free from any of the ICM-MM conditions by age 50 was below 40%. For a 30-year-old Bangladeshi woman, the probability of remaining free from any of the ICM-MM conditions by the age of 40 was below 50%.

**FIG 4a.**
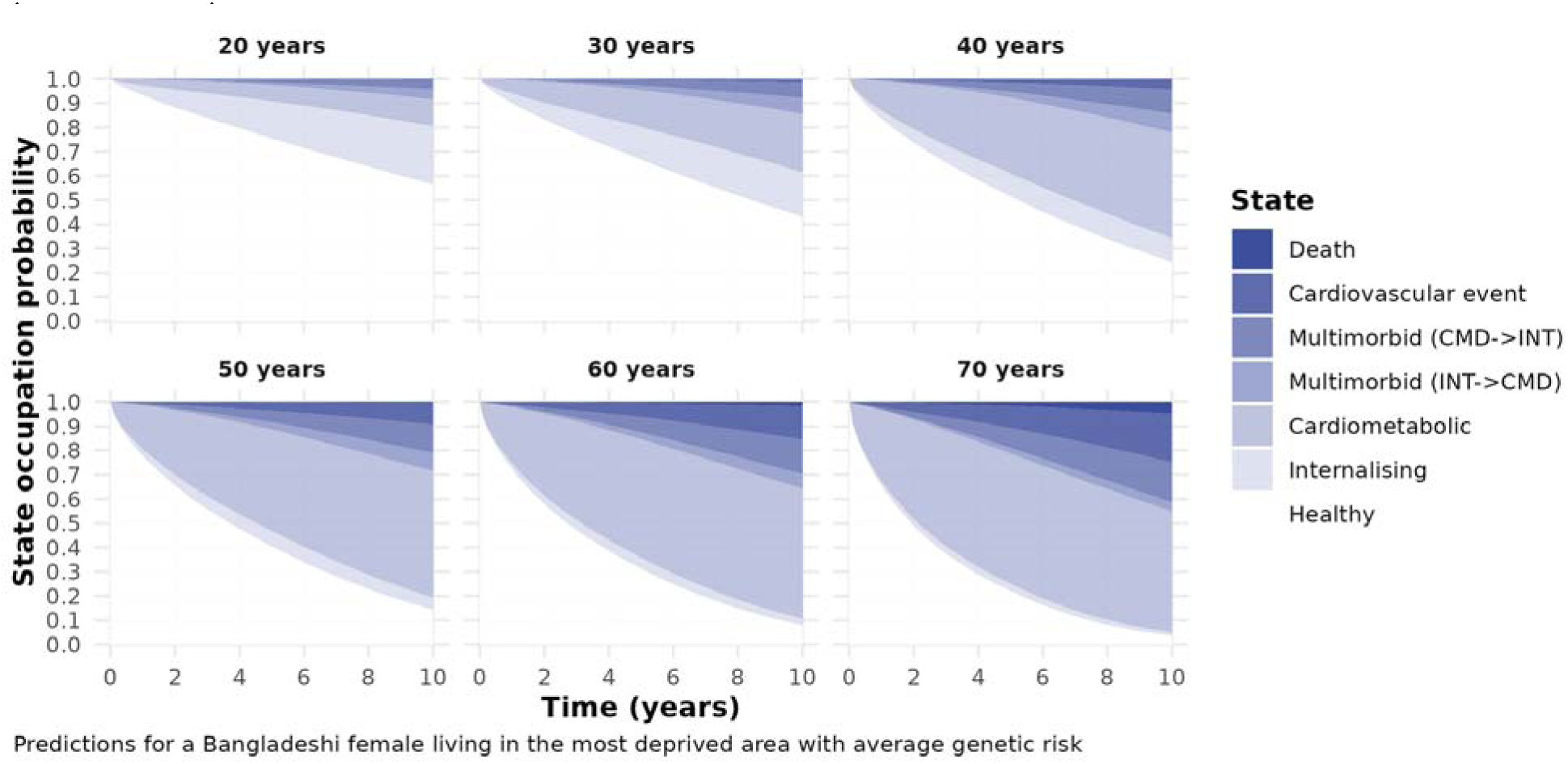
Predicted health state occupation by baseline age

**FIG 4b.**
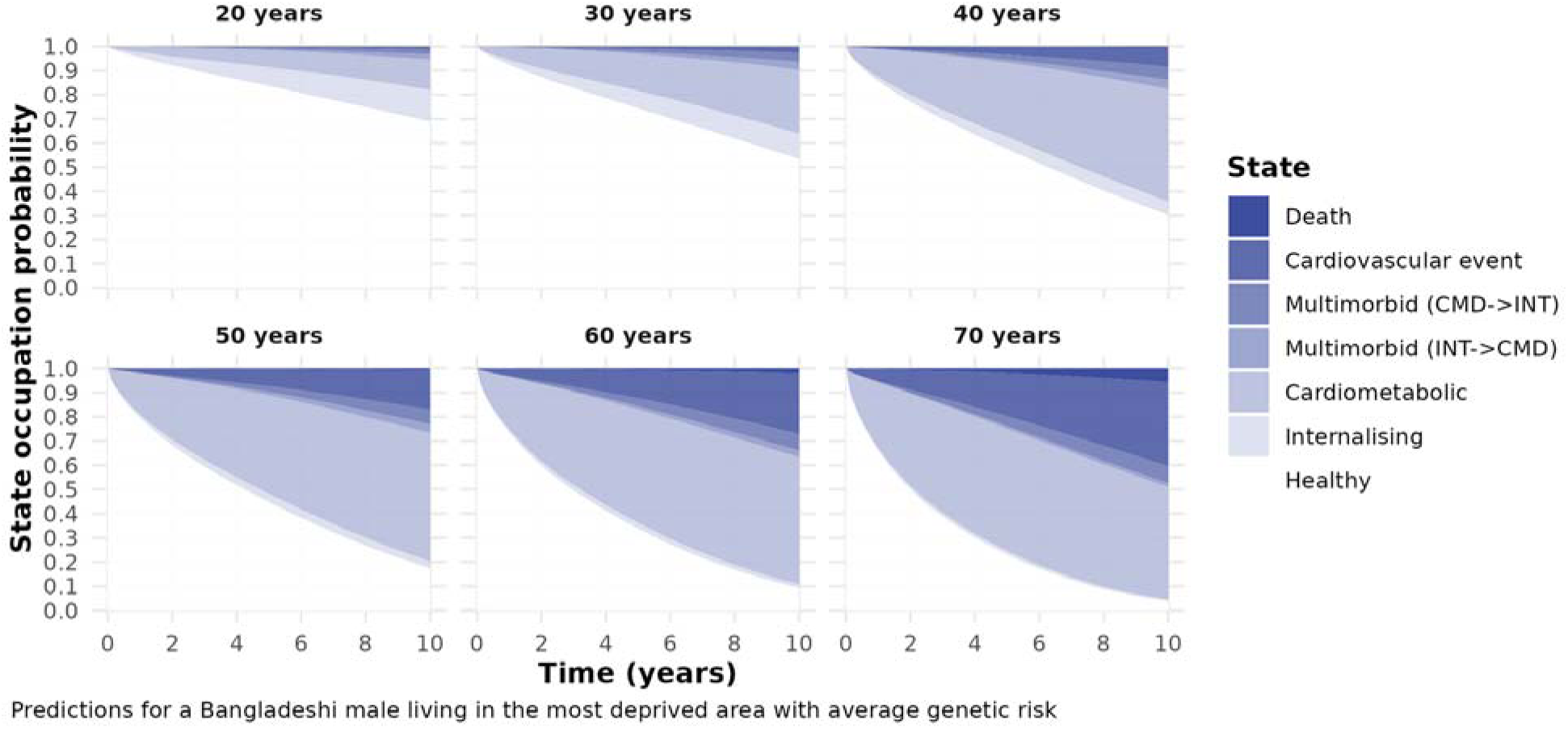
Predicted health state occupation by baseline age (males)

We also observed a non-proportional hazard for baseline age in transitions to internalising states suggesting that people were more likely to report internalising conditions at younger ages. Across all age groups the probability of remaining healthy decreased over the ten-year period. This was especially apparent in older age groups, where the proportion of people likely to experience cardiometabolic conditions in particular was very high. Even in younger age groups (20 and 30) the probability of experiencing multimorbidity over a ten-year time frame was between 5% and 10%. In the youngest age groups (20 and 30), multimorbid states were equally probable via cardiometabolic → internalising and internalising → cardiometabolic. From 40 onwards, multimorbidity was more likely via cardiometabolic→internalising, and the probability of experiencing death or CVE over a ten-year timeframe increased greatly after the age of 50.

#### Condition ordering and CVE risk

The 10-year probability of CVE was higher for cardiometabolic→ internalising across all baseline ages tested, and in women and men (SUPPLEMENTAL FIGURES 1 and 2), but the difference was only conventionally statistically significant for women who developed ICM-MM at the age of 40 *P(CVE | internalising* → *cardiometabolic)* = 0.056 vs *P(CVE| cardiometabolic* → *internalising)* =0.116, *contrast(P)*=0.058, 95%CI:0.017 – 0.100. The equivalent contrast for men was *P(CVE | internalising* → *cardiometabolic)* = 0.095 vs *P(CVE| cardiometabolic*→*internalising)=0.161*, *contrast(P)*=0.078, 95%CI:-0.00 – 0.140

### Sociodemographic and genetic contrasts

#### Gender

[<u>FIGURE5</u>] shows the difference in probabilities for being in each health state for a man, versus the reference woman over a ten-year window and associated confidence intervals. Men were more likely to remain free of ICM-MM conditions (‘otherwise healthy’), and less likely to experience internalising states (and hence ICM-MM, either via *internalising* → *cardiometabolic* or *cardiometabolic*→*internalising*). However, men had a higher probability of experiencing a CVE over the 10-year window.

**FIG 5.**
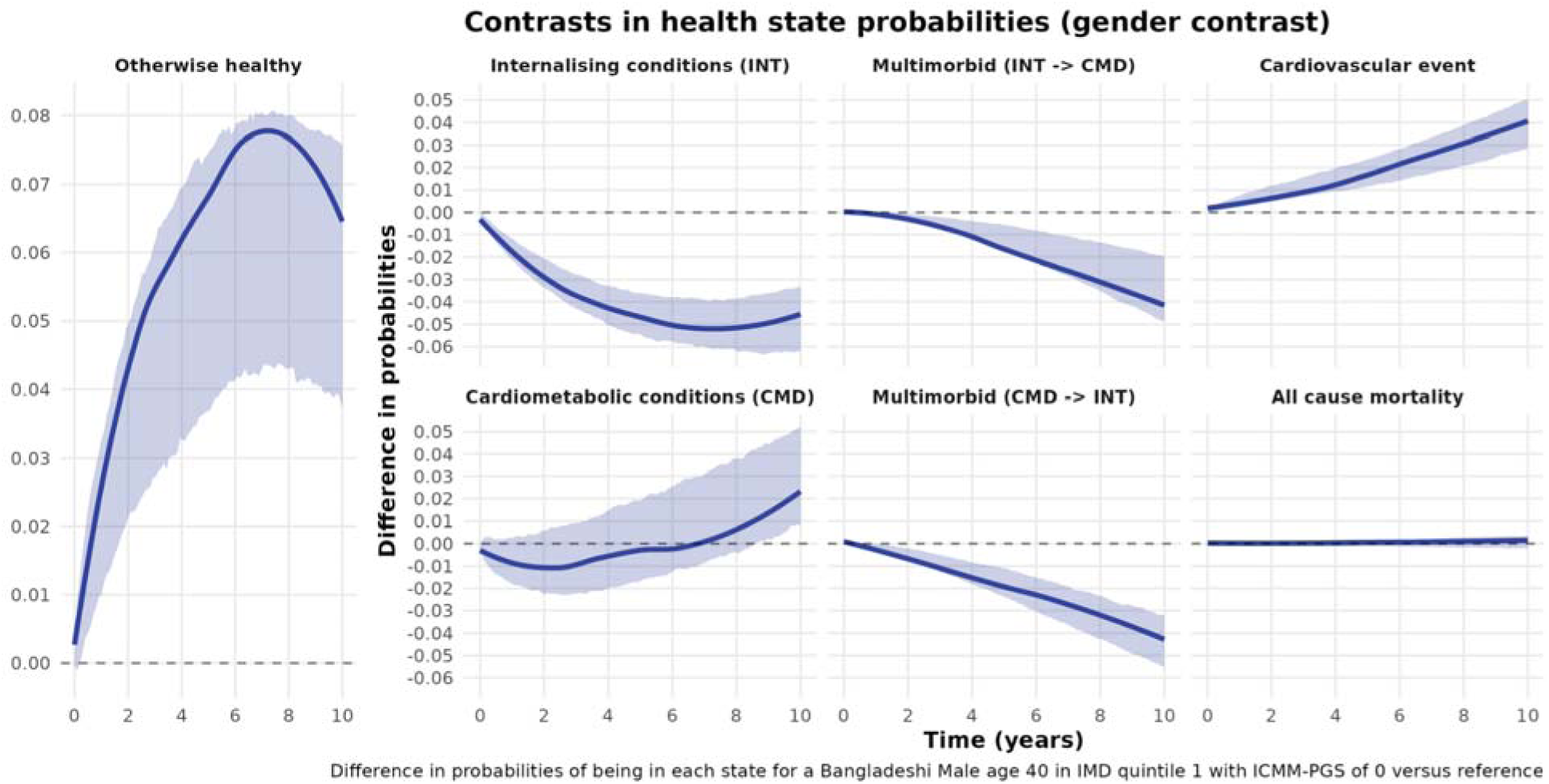
Contrast - gender

#### Ethnicity

[<u>FIGURE6</u>] shows the difference in probabilities of health state occupation for a Pakistani woman versus the reference individual. The probability of remaining otherwise healthy (free from ICM-MM conditions) remains higher over the ten-year window for the Pakistani individual - primarily due to the reduced probability of developing a cardiometabolic condition. However, the difference in probabilities of developing ICM-MM is only slightly lower for the Pakistani individual for both multimorbid states, particularly for *internalising* → *cardiometabolic*.

**FIG 6.**
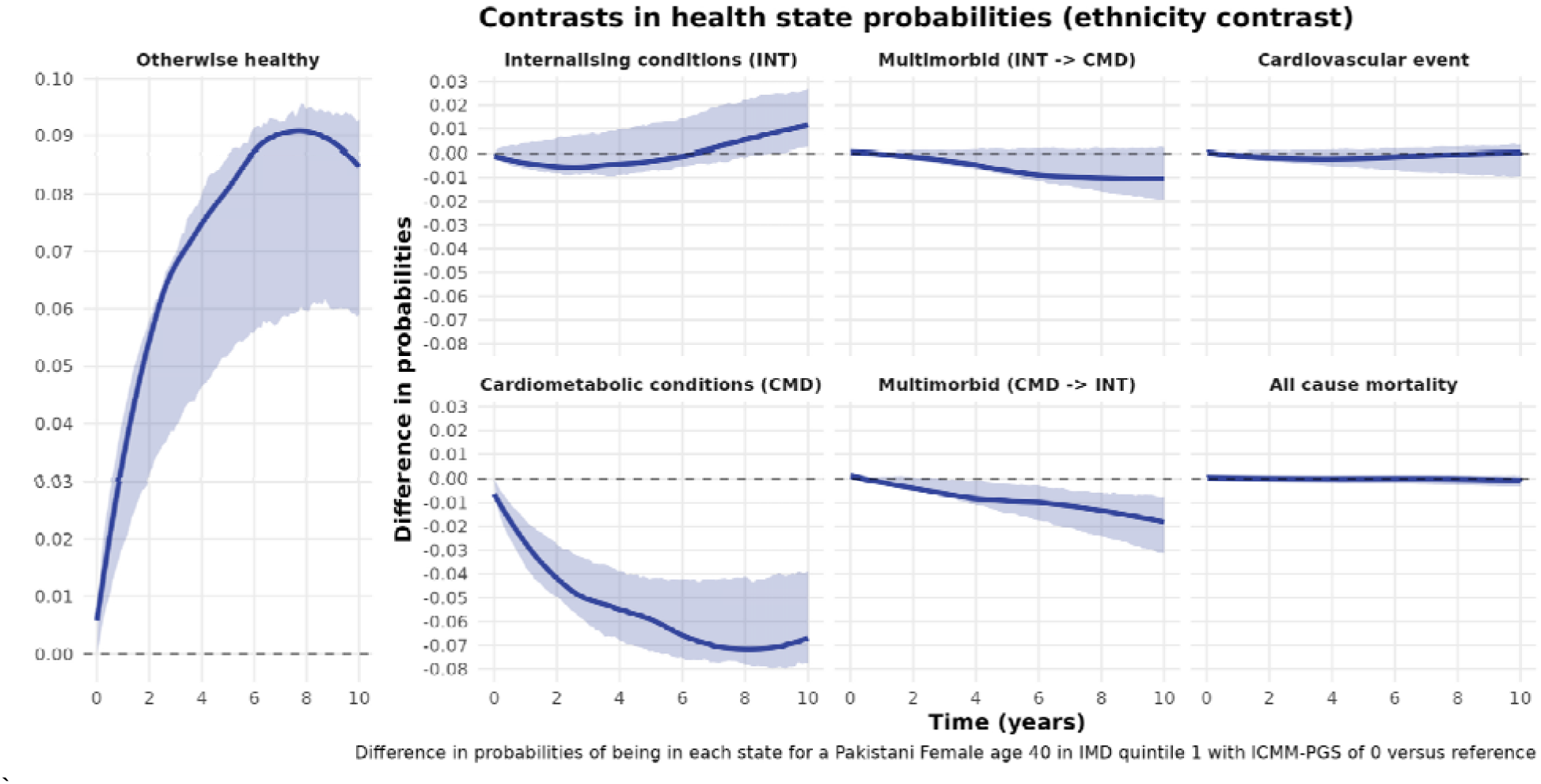
Contrast - Ethnicity

#### IMD

[<u>FIGURE7</u>] shows the difference in probabilities of health state occupation for someone living in the least deprived tertile (IMD 3+) versus the reference (IMD 1). The probability of remaining healthy was higher for the individual living in the least deprived tertile. Differences in overall health are driven by a reduced probability of experiencing all of the internalising, cardiometabolic and ICM-MM health states conditions, though there was no difference in the probability of experiencing CVE or death.

**FIG 7.**
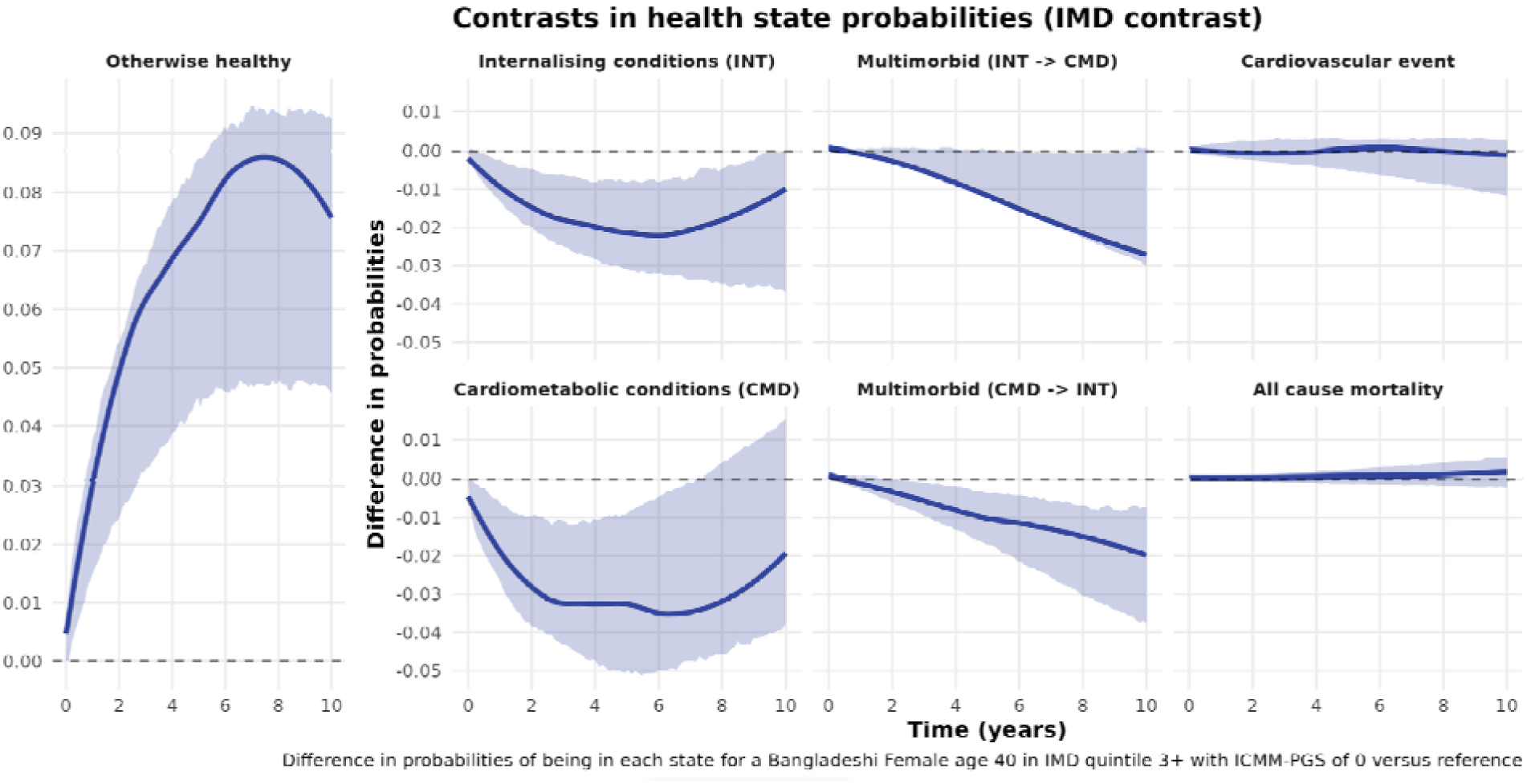
Contrast - IMD

#### Smoking

[<u>FIGURE8</u>] shows the contrast for an ever vs never smoker at study baseline. The probability of an ever smoker remaining free from ICM-MM conditions was up to 3% lower over the 10-year window versus never smokers. Ever smoking was also associated with increased probability of internalising conditions as the index condition, and increased ICM-MM risk along both trajectories (*cardiometabolic* → *internalising* and *internalising* → *cardiometabolic*) following the index condition. The probability of CVE was also higher over the 10-year window.

**FIG 8.**
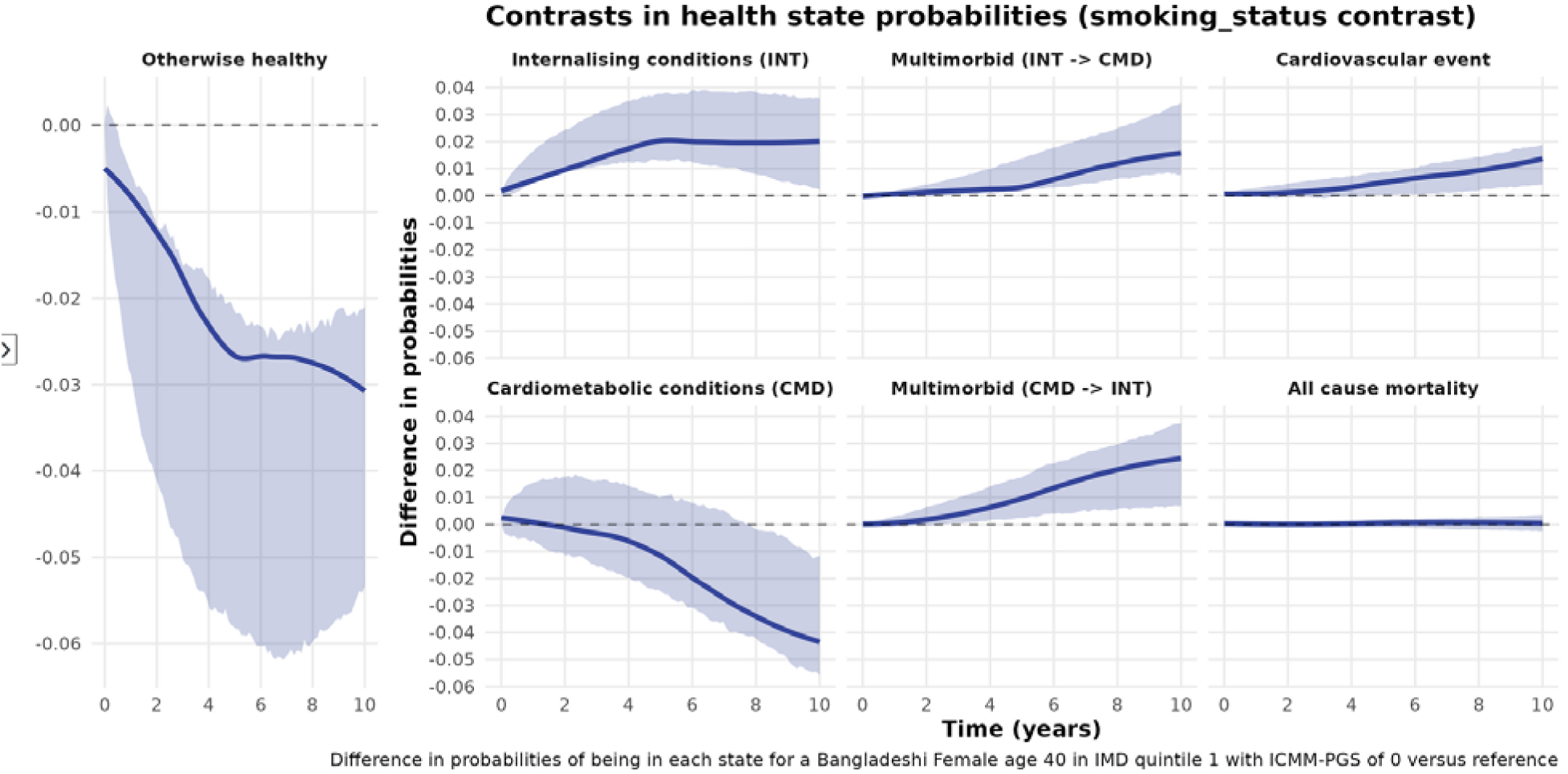
Contrast - Smoking

#### ICM-MM_PRS_

[<u>FIGURE9</u>] shows the contrast in state occupation probabilities for someone in the top 2.5% of polygenic risk (ICM-MM_PRS_ z-score = 2) vs someone with average genetic risk (ICM-MM_PRS_ z-score= 0). Participants with a higher ICM-MM_PRS_ had a higher probability of experiencing cardiometabolic → internalising (but not internalising→cardiometabolic) trajectories and higher probability of CVE.

**FIG 9.**
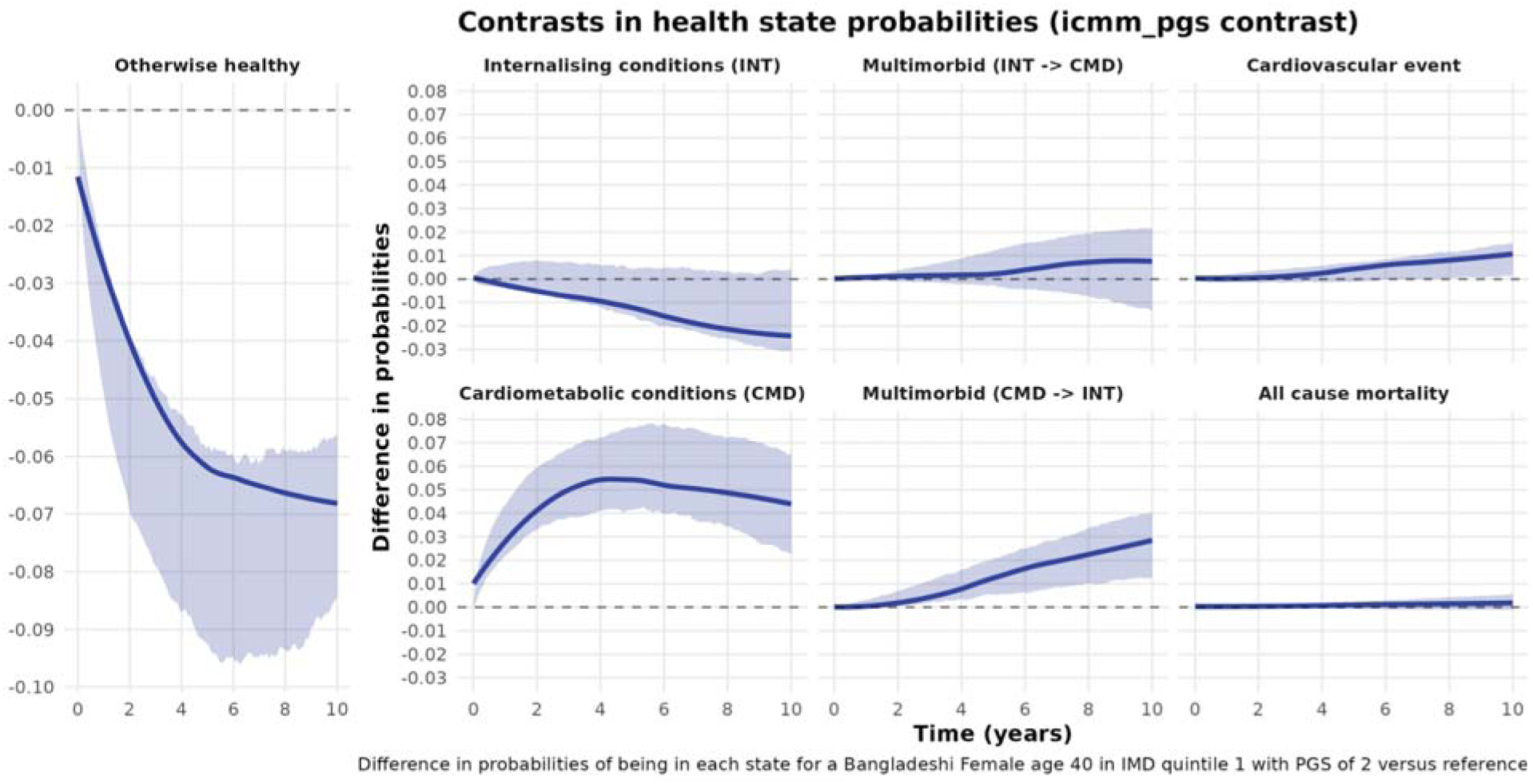
Contrast - ICMM-PRS

## Discussion

In this study, we examined internalising and cardiometabolic multimorbidity trajectories and risk of cardiovascular events and death in a large cohort of British Pakistani and British Bangladeshi participants. Combining real world healthcare data with genetic information, we examined the impact of sociodemographic and genetic factors on these multimorbidity trajectories. We found rapid accumulation of illness and multimorbidity, even in young age groups. Assuming good health at the age of 40, the probability that Bangladeshi participants living in the most deprived areas will remain free from ICM-MM conditions over the next ten years is below 40%. For a healthy Bangladeshi woman aged 30, the probability that she will remain free of ICM-MM conditions by age 40 is below 50%. In the UK, individuals are eligible for routine screening as part of the nationwide NHS health check from age 40; our findings suggest that specific populations, such as Bangladeshi women, may benefit from earlier initiation of routine health screening with the potential for improved public health outcomes.

We found evidence that people who develop ICM-MM along the trajectory cardiometabolic→internalising have a higher probability of developing a cardiovascular event (CVE) over the next ten years than the internalising→cardiometabolic trajectory, though the evidence for this was strongest for women who developed ICM-MM at age 40. This finding builds on an established literature showing that individuals with type 2 diabetes are more likely to have a CVE if they have co-existent depression.^50^ However, the lack of longitudinal data in these prior studies means it is not possible to see whether this association may be driven by depression prior to, or after, the diagnosis of type 2 diabetes. By characterising the ordering of conditions, we can hypothesise why there are differences in progression to CVE events. Individuals who develop internalising conditions after cardiometabolic disease may be experiencing a greater treatment burden from that cardiometabolic disease, leading to depression and/or anxiety, or may be non-adherent to medications that offer protection against CVE.^51^ In contrast, individuals who have established internalising conditions at the time of diagnosis of a cardiometabolic disease may receive holistic care that addresses both, e.g. in the UK, screening and management of depression and anxiety is encouraged at type 2 diabetes diagnosis,^52^ and structured education programmes delivered to people with type 2 diabetes, typically at or soon after diagnosis, have well-characterised psychosocial benefits.^53^

Our findings also build on existing evidence of the shared genetic architecture of many cardiometabolic and mental health traits, ^54–56^ demonstrating that this architecture has consequences for ordering of condition pairs. The polygenic risk score developed to predict ICM-MM (ICM-MM_PRS_), was associated with higher risk of ICM-MM via the trajectory cardiometabolic→ internalising (the trajectory resulting in the highest probability of CVE) than for the trajectory internalising→cardiometabolic. This earlier onset cardiometabolic profile with higher internalising condition risk (and subsequent risk of cardiovascular event) should be a target for early intervention.

We also found that younger otherwise healthy participants were more likely to transition to internalising states (in any order) than older participants. This finding is in line with large scale studies on increasing prevalence of depression in the UK, especially in younger cohorts.^57^ Our findings may have implications on future ICM-MM prevalence in this cohort - especially under the ‘lifetime occurrence’ definition of multimorbidity^3^ - as younger participants continue to age. This observation may also reflect changes in behaviour and attitudes towards depression in South Asian populations,^58^, as well as improved ascertainment by primary care clinicians.^59^

We also found that Bangladeshi participants were more likely than Pakistani participants to experience ICM-MM conditions, ICM-MM and CVE, highlighting a health inequality previous large-scale registry based studies on multimorbidity trajectories in the UK would not have been able to detect due to aggregation across ethnicity groupings in the case of Chen et al., 2023,^13^ and absence of information on ethnicity in Owen et al (2023)^11^ and Lyons et al (2023).^12^ Ethnicity is a complex multidimensional construct that encapsulates cultural, religious and many other factors.^14^ Smoking is one important potential source of variation associated with ethnicity ^60^ - and Bangladeshi participants were more likely to be ‘ever’ smokers than Pakistani participants in our sample. We have adjusted for smoking in our analysis, but future work should investigate other environmental, sociodemographic and lifestyle factors that may be responsible for the differences we observed.

We observed similar associations between area level deprivation (IMD) and disease transitions to those found in previous multimorbidity studies.^5^ ^13,61^ Participants living in the most deprived quintile were more likely to experience internalising and cardiometabolic conditions, ICM-MM and CVE than those living in three least deprived quintiles. However, most G&H participants live in areas that are in the two most deprived IMD quintiles, concentrated in East London, meaning many participants are likely to have similar exposure to environmental factors that are known to be associated with multimorbidity - air pollution in particular ^62^ - but that are not well reflected in the aggregated IMD.

### Limitations

Our study uses routinely collected electronic healthcare records, the limitations of which are well reported (see Farmer et al for an example).^63^ We adjusted for area-level deprivation (IMD), which is a commonly used metric to assess environmental risk exposures in England, but we note this is subject to the ecological fallacy).^64^ As this was ascertained at study baseline, any changes in living circumstances (and their impact on health trajectories over the course of follow-up) are not reflected in our results. This is also the case for our ascertainment of ‘ever’ vs ‘never’ smoking status - smoking cessation is likely to reduce or delay onset of cardiometabolic multimorbidity.^55^ In common with other studies that have used electronic healthcare records to define health phenotypes we have made a series of assumptions about ascertainment and chronicity of the conditions, as there is often no triggering contact that would make a clinician record remission/recovery. Using diagnosed obesity as part of the cardiometabolic condition definition also removed our ability to investigate BMI as a potential moderator on the proposed causal relationship between depression and diabetes.^65^

One limitation of our novel ICM-MM_PRS_ is the lack of cross-ancestry GWAS available to create the contributing PRS, which are drawn from predominantly white European ancestry populations. Previous work has demonstrated the power of cross-and multi-ancestry genetic instruments to predict cardiometabolic traits^66^ and further work is needed to improve our understanding of cross-ancestry and ancestry specific genetic risk for multimorbidity. We also note that a recent study of a more restricted physical-mental health multimorbidity cluster,^67^ reported a PRS with similar performance to ICM-MM_PRS_, which suggests we are near the ceiling for prediction of heterogeneous multimorbidity clusters via polygenic risk scores rather than a limitation of our instrument *per se*.

## Conclusions

Our work highlights the high burden of morbidity and multimorbidity in UK Pakistani and Bangladeshi populations, with clear social and demographic patterning of physical and mental health morbidity. The early onset of many of the ICM-MM conditions, and ICM-MM itself suggests earlier intervention and screening may be required in these populations to reduce the impact of living with multiple long-term conditions, and future CVE risk. Constructing genetic risk models for physical and mental health multimorbidity has only a modest impact on determining risk but may be a feasible addition to future precision-based care models using genotypic data.

We demonstrate the utility of taking a lifecourse approach to studying the development of multimorbidity, observing patterns such as the emergency of ICM-MM in young Bangladeshi women following a single internalising condition. This observation highlights the need for holistic care models that proactively screen for and intervene early in people with internalising conditions. Future research could use trial emulation within existing models of clinical care to determine whether the progression between states of multimorbidity, and to cardiovascular disease and death can be prevented.

## Data Availability

All data produced in the present study are available upon reasonable request to the authors

https://www.genesandhealth.org/researchers/apply-for-access/

## Acknowledgements

We would like to thank Chris Jackson, author of the flexsurv package for his helpful correspondence on implementing and updating some of flexsurv’s features, and for continuing to support the flexsurv package.

## LINC

This work was funded by the Tackling Multimorbidity at Scale Strategic Priorities Fund programme [grant number MR/W014416/1] delivered by the Medical Research Council and the National Institute for Health Research in partnership with the Economic and Social Research Council and in collaboration with the Engineering and Physical Sciences Research Council. For further details of the LINC project, please visit our website: https://.uk/lifespan-multimorbidity-research-collaborative

## Genes & Health

Genes & Health is/has recently been core-funded by Wellcome (WT102627, WT210561), the Medical Research Council (UK) (M009017, MR/X009777/1, MR/X009920/1), Higher Education Funding Council for England Catalyst, Barts Charity (845/1796), Health Data Research UK (for London substantive site), and research delivery support from the NHS National Institute for Health Research Clinical Research Network (North Thames). Genes & Health is/has recently been funded by Alnylam Pharmaceuticals, Genomics PLC; and a Life Sciences Industry Consortium of Astra Zeneca PLC, Bristol-Myers Squibb Company, GlaxoSmithKline Research and Development Limited, Maze Therapeutics Inc, Merck Sharp & Dohme LLC, Novo Nordisk A/S, Pfizer Inc, Takeda Development Centre Americas Inc.

We thank Social Action for Health, Centre of The Cell, members of our Community Advisory Group, and staff who have recruited and collected data from volunteers. We thank the NIHR National Biosample Centre (UK Biocentre), the Social Genetic & Developmental Psychiatry Centre (King’s College London), Wellcome Sanger Institute, and Broad Institute for sample processing, genotyping, sequencing and variant annotation.

## Data sharing

Access to anonymised clinical data and genotyping used for all analyses, figures, and tables are available to the research community via a Trusted Research Environment upon approval by the Genes & Health scientific committee and the independent Information Governance Review Panel. All codelists used to define health phenotypes are publicly available. All code used in the analyses are available from the authors on request.

